# Factors associated with chemotherapy-induced peripheral neuropathy-related reduced taxane dose or premature discontinuation in women with early-stage breast cancer

**DOI:** 10.1101/2021.09.13.21263539

**Authors:** Lynn R. Gauthier, Lye-Ann Robichaud, Maud Bouffard, Frédérique Therrien, Sarah Béland, Marianne Bouvrette, Jennifer Gewandter, Lucia Gagliese, Robert H Dworkin, Julie Lemieux, Josée Savard, Philip L. Jackson, Michèle Aubin, Sophie Lauzier, Bruno Gagnon, Anne Dionne, Cindy Shobbrook, Pierre Gagnon

**Affiliations:** CHU de Québec-Université Laval Research Center, Oncology Division, Quebec, Canada; School of Psychology, Faculty of Social Sciences, Université Laval, Quebec, Canada; Department of Psychology, Faculty of Arts and Science, Université de Montréal, Montreal, Canada; Department of Family and Emergency Medicine, Faculty of Medicine, Université Laval, Quebec, Canada; Department of Community Health Sciences, Faculty of Medicine and Health Sciences, Université de Sherbrooke, Sherbrooke, Canada; Université Laval Cancer Research Center, Quebec, Canada; Department of Anesthesiology and Perioperative Medicine, University of Rochester Medical Center School of Medicine and Dentistry, Rochester, United States of America; Department of Neurology, University of Rochester Medical Center School of Medicine and Dentistry, Rochester, United States of America; School of Kinesiology and Health Science, York University, Toronto, Canada; Departments of Anesthesia and Psychiatry, University of Toronto, Toronto, Canada; Centre des maladies du sein Deschênes-Fabia, CHU de Québec, Quebec, Canada; Centre interdisciplinaire de Recherche en Réadaptation et Intégration Sociale (Cirris),, Quebec, Canada; CERVO Research Center, Québec, Canada; Waterloo Wellington Integrated Hospice Palliative Care Program, Waterloo, Canada; Faculty of Pharmacy, Université Laval, Quebec, Canada; CHU de Québec-Université Laval Research Center, Population Health and Optimal Health Practices Division, Quebec, Canada; Department of Psychiatry and Neurosciences, Université Laval, Quebec, Canada

## Abstract

**Purpose:** In the absence of treatments for chemotherapy-induced peripheral neuropathy (CIPN), dose reductions (DR) and premature discontinuation (PD) are primary management strategies. However, decision-making guidance is insufficient and knowledge of factors associated with DR/PD is limited. We examined biopsychosocial factors associated with CIPN-related DR/PD in women undergoing taxane-based chemotherapy for early-stage breast cancer.

**Patients and methods:** As part of a longitudinal study of CIPN measurement, women completed assessments before the first taxane infusion and at the final infusion or within the originally expected timeframe for the final infusion. Participants completed self-report measures of CIPN, pain, and physical and psychosocial wellbeing, and underwent physical testing of lower limb disability and Quantitative Sensory Testing for sensation and pain threshold to thermal, vibration, and touch stimuli in the feet and hands. Sociodemographic and clinical data were collected. Logistic regression was used to identify factors associated with neuropathy-related DR/PD.

**Results:** Among 121 participants, 66 (54.5%) received taxane-as-prescribed, 46 (38.0%) had neuropathy-related DR/PD, and 9 (7.4%) had DR/PD for other reasons. Factors associated with neuropathy-related DR/PD were receipt of paclitaxel (Odds Ratio [OR]=75.05, 95% Confidence Interval [CI] 2.56-2197.96]), lower pre-treatment pain catastrophizing (OR=0.72, 95% CI: 0.54 – 0.95), and higher post-treatment neuropathic pain (OR=10.77, 95% CI: 1.99 – 58.15) and sensitivity to cold pain in the hand (OR=1.64, 95% CI: 1.05 – 2.56).

**Conclusion:** CIPN-related DR/PD is associated with paclitaxel treatment and post-treatment neuropathic pain and cold pain sensitivity in the hands. CIPN communication to healthcare providers may be influenced by pain catastrophizing, suggesting symptom appraisal may be an important factor in communication. Findings could contribute to clinical practice recommendations to facilitate treatment decision-making.

**Lay summary:** We studied pre- and post-treatment factors associated with reduced taxane dose or early cessation due to chemotherapy-induced peripheral neuropathy in women undergoing chemotherapy for early-stage breast cancer. Reduced taxane dose or early cessation is associated with paclitaxel treatment, and high post-treatment neuropathic pain and sensitivity to cold pain stimuli in the hands. Communication of these experiences to healthcare providers may be influenced by pre-treatment thoughts and feelings about symptoms.

**Precis for use in the Table of Contents:** two concise sentences that state the significant conclusion(s) or message of the manuscript; Chemotherapy-induced peripheral neuropathy-related reduced taxane dose or premature discontinuation is associated with paclitaxel treatment and high post-treatment neuropathic pain and cold pain sensitivity in the upper limbs. Reporting of these experiences may be influenced by pre-treatment symptom appraisal and communication style.

## INTRODUCTION

Chemotherapy-induced peripheral neuropathy (CIPN) affects 43%–80% of women undergoing taxane-based treatment (paclitaxel, docetaxel) for breast cancer (BC)^1–3^. It involves sensory symptoms like pain, numbness, tingling, and/or cold allodynia, with motor weakness occurring at high doses^4,5^. Duloxetine is the only treatment recommended for painful CIPN^4,6^. However, its efficacy for the myriad sensory, motor, and autonomic symptoms other than pain has not been established.

In the absence of available treatments, dose reductions or premature discontinuation (DR/PD) are primary management strategies^7^, which have been reported in 5% to 35% of women with early-stage BC^3,10–15^. The American Society of Clinical Oncology (ASCO) guidelines recommend discussion with patients about treatment changes, including DR/PD, “in the case of intolerable neuropathy and/or functional nerve impairment”^4^. However, they offer no guidance about the characteristics of intolerance or impairment warranting a DR/PD decision. Similarly, clinical monograph treatment change recommendations are inconsistent and insufficient for decision-making^14^.

CIPN-related DR/PD risk factor knowledge is limited. While paclitaxel is associated with CIPN-related treatment alterations, tumor characteristics, age, alcohol use, and history of diabetes are not^12,25^. Unfortunately, no studies have compared people with CIPN-related DR/PD to people who received treatment-as-prescribed on a broad range of biopsychosocial factors. This could provide important information about potentially modifiable factors.

Like other cancer pain^26,27^, CIPN is associated with biopsychosocial factors. Since the guidelines recommend DR/PD discussion for intolerable CIPN^4^, an understanding of biopsychosocial factors associated with CIPN presence or severity may provide insight. CIPN presence and severity are associated with paclitaxel treatment and dose^18^, greater post-treatment depression, anxiety, and insomnia, lower quality of life (QOL)^19–21^ and physical wellbeing^3^, balance problems and falls^29,32–34^, and impaired functioning^35,36^. Greater pre-treatment anxiety, depression and fatigue also predict post-treatment CIPN presence and severity^37,38^.

Since the guidelines also recommend DR/PD discussion for impaired nerve fiber functioning^4^, quantitative sensory testing (QST) may also provide insight. QST quantifies sensory nerve fiber functioning through tests of sensation and pain threshold to stimuli, like thermal, touch, and vibration. It can track sensory changes throughout treatment^17,18,39^, improve CIPN detection, and guide decision-making^7,40^. For example, cold allodynia and hyperalgesia in the hand predicts severe neuropathy^33^, people with CIPN have lower heat pain threshold than people without CIPN^34^, and moderate-to-severe CIPN is associated with reduced tactile and vibration perception in some studies^26^, but not others^35^. Studies of CIPN-related DR/PD using QST do not exist. Thus, the role of impaired nerve fiber functioning is unknown.

Insufficient decision-making guidance is problematic: By the time DR/PD must be considered, CIPN reversibility is unknown. Moreover, DR/PD may reduce treatment efficacy, with unclear impacts on long-term survival^14^. A better understanding of factors associated with CIPN-related DR/PD could improve decision-making guidance by clarifying the type of intolerance and functional nerve impairment associated with DR/PD and identifying people at-risk to intervene on modifiable factors. This study aims to evaluate biopsychosocial factors associated with CIPN-related DR/PD in women undergoing taxane-based chemotherapy for early-stage BC.

## PATIENTS AND METHODS

### Study Design, Setting and Participants

As part of a longitudinal study on CIPN measurement, women with early-stage BC were recruited before starting taxane-based chemotherapy at the Centre des maladies du sein Deschênes-Fabia, CHU de Québec-Université Laval in Quebec, Canada between February 2017 and May 2019. Eligible patients were ≥18 years old, able to read, write and understand French, and chemotherapy naïve. Exclusion criteria were metastatic disease, comorbidities associated with peripheral sensory abnormalities (e.g., diabetes, HIV/AIDS, alcohol abuse [NIH criteria^36^], polyneuropathy) and scores <20 on the Short Orientation Memory Concentration Test^37^, a brief cognitive screen. Approval was received from the Research Ethics Committee of the CHU de Québec-Université Laval.

The project was presented during weekly chemotherapy patient education classes. Interested attendees signed a pre-consent form granting permission to access their medical files to verify eligibility and be contacted via telephone. Study visits were scheduled before the first chemotherapy cycle or ≤2 weeks for regimens not starting with taxane (T0). Written informed consent was obtained before data collection. The second assessment (T1) took place at the final taxane infusion (±1 week) or within the originally expected timeframe for the final infusion^38^.

At each assessment, sociodemographic and clinical information were collected or updated and a short QST protocol^39^ and a physical functioning assessment were conducted. Self-report measures were completed before the first infusion (T0) and ≤7 days after T1. Questionnaire, QST, and physical functioning test order was randomized. Clinical data were abstracted from the electronic medical record (EMR) at T0 and updated at T1.

### Measures

Sociodemographic data, and pain history, and management were collected via brief interview. Clinical data abstracted from the EMR and the CHU de Québec-Université Laval oncology registry included information about cancer and treatment factors, medical comorbidities, and prescribed medications. The Research Assistant (RA) completed the **Charlson Comorbidity Index (CCI)**^40^ and calculated the **Anticholinergic Drug Scale** (**ADS**)^41^ because of the relationship between anticholinergic load and pain^42^.

#### Outcome variable

Following chart review, participants were categorized into 3 groups according to receipt of prescribed taxane and reasons for DR/PD. DR/PD was coded as neuropathy-related if consultation notes mentioned neuropathy, paresthesia, dysesthesia, pain in the periphery, and/or motor weakness as the reason for DR/PD. Each participant was coded by two independent raters. Disagreements were reviewed until consensus was reached. Group 1 (G1) received taxane-as-prescribed, Group 2 (G2) had neuropathy-related DR/PD, and Group 3 (G3) had DR/PD for other reasons.

#### Pain, fatigue and sleep, psychosocial wellbeing, and quality of life self-report measures

The **Pain History Questionnaire (PHQ**)^43^ collected information about the presence of cancer-related and non-cancer pain on most days in the past 3 months. We also calculated average expected pain immediately and 1 week after chemotherapy, and after pain treatment on 0 (*no pain*) to 10 (*worst possible pain*) numeric rating scales. Pain Severity and Interference were measured with the **Brief Pain Inventory (BPI)**^44^. Non-neuropathic, Neuropathic and Affective pain qualities were measured with the **Short-Form McGill Pain Questionnaire-2 (SF-MPQ-2)**^45^. Spontaneous ongoing pain, paroxysmal and evoked pain, dysesthesia and paresthesia were measured with the **Neuropathic Pain Symptom Inventory (NPSI)**^46^. CIPN-related neurotoxicity symptoms, motor weakness, arthralgia, myalgia, and skin changes were measured with the **Functional Assessment of Cancer Therapy-Taxane subscale (FACT-Taxane)**^47^.

Sleep quality and cancer-related fatigue were measured with the **Pittsburgh Sleep Quality Index (PSQI)**^48^ and the **Functional Assessment of Chronic Illness Therapy (FACIT)-Fatigue Scale**^49^. Cancer-related physical, social/family, emotional, and functional QOL were measured with the **FACT-General**^47^. Depressive symptoms were measured with the **Center for Epidemiologic Studies–Depression Scale (CES-D**)^50^. Rumination and catastrophic negative thoughts about pain and its management were measured with the **Pain Catastrophizing Scale (PCS)**^51^. All measures have been validated for use in cancer or pain. Higher FACIT-Fatigue, and FACT-Taxane and -General, reflect lower fatigue and neurotoxicity, and better QOL. Higher scores on other measures reflect greater pain, symptoms, or worse wellbeing.

#### Quantitative sensory testing

Using an adapted protocol^39^ we assessed cold (CPT) and heat pain threshold (HPT), and cool (CDT), warm (WDT), vibration (VDT; TSAII NeuroSensory Analyzer & VSA 3000, Medoc, Israel), and mechanical detection threshold (MDT; SenseLab Aesthesiometer II hand-held nylon filaments, Somedic, Sweden) on the dorsum of the right hand and foot^41,59^. A 3cm × 3cm thermode was used for thermal tests. Baseline temperature was 32°C with 0.5°C/sec rate of change and maximum cooling and warming at 0° C and 50.5°C. The thermode was immediately removed upon response. Baseline vibration was 0μm/sec with 1μm/sec rate of change. Thermal and vibration thresholds were calculated from the mean of three trials. MDT was calculated as the geometric mean (grams). Higher WTD, HPT, VDT, and TDT reflect less sensitivity. Higher CDT and CPT reflect greater sensitivity.

#### Physical functioning

The **Karnofsky Performance Status Scale (KPS)**^60,61^, a measure of observer-rated functioning, was completed by the RA. Use of a **walking accessory** and **number of falls**^24^ or near-falls in the past week (T0) and since T0 (T1) were recorded by the RA. Balance and mobility was assessed with the **Timed Up and Go Test (TUG)**^62,63^ which was calculated from the mean of 2 trials. The **Short Physical Performance Battery Protocol (SPPB)**^57^ was performed to assess static balance.

### Analyses

Missing data were screened and the pattern analyzed using Little’s Missing Completely at Random test^58^. Where self-report items were missing-at-random or completely-at-random, scores were calculated based on completed items. Scores were not imputed for measures missing data on all items. Statistical significance was set at p≤0.05. T-tests and χ^2^ analyses assessed whether participants who completed both assessments differed from those who did not complete T1 on baseline data. Means, standard deviations (SD), frequencies, and percentages were used to characterize participants. Repeated measures t-tests, Cochrane Q, and Wilcoxon tests were used to evaluate change.

To determine candidate correlates for the multivariate regression model, bivariate analyses, including one-way ANOVAs and χ^2^ analyses were used to compare groups on sociodemographic, clinical, and T0 and T1 QST, physical functioning, and self-report measures. Post-hoc analyses determined pair-wise group differences. Correlates significantly different (p≤0.05) across groups were considered for model entry, a more conservative level than recommended^59^ to control for overfitting. We included corresponding T0 variables for each T1 variable included to control for pre-treatment values, and vice versa. Correlates highly associated with each other (r≥0.7)^59,60^ were evaluated for missing data, assumption violations, or effect sizes to determine which variable to retain. We used standard binomial logistic regression as G3 was too small taking into account missing data (n=4). Analyses were conducted using SPSSv25.

## RESULTS

Figure 1 describes participant recruitment and retention. 154 (59,2%) women consented to participate. Five (3.2%) were subsequently excluded and 1 (0.6%) withdrew before T0. 148 and 121 (81.1% retention) participants completed T0 and T1, respectively. Compared to participants who completed T1, those who did not were older (*p*≤0.05), less educated (*p*≤0.001), less likely to exercise (*p*≤0.05), had higher BMI (*p*≤0.05) and KPS (*p*≤0.05), and were less sensitive to foot and hand MDT (*p*≤0.05). Data were missing at random^58^ and were not associated with baseline self-report pain or symptoms.

**Figure 1.**
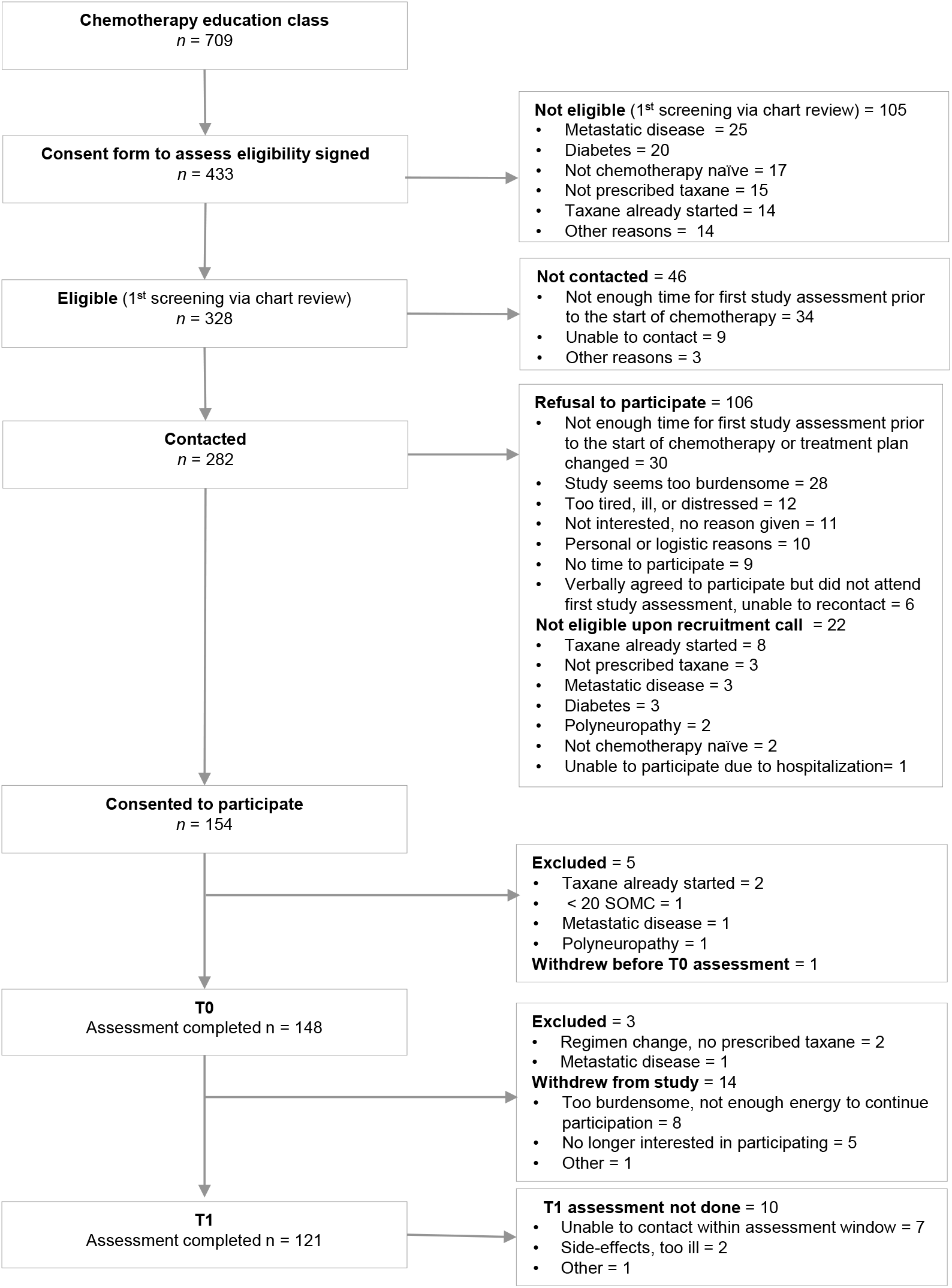
Flowchart summarizing participation, main exclusion and withdrawal reasons

### Participant characteristics

Subsequent analyses include participants who completed both assessments (n=121). Descriptive data and T0-T1 change are presented in Table 1. Participants were 50.9±11.1 years old, mostly Caucasian, partnered, and well-educated. Few (2.5%) had ≥1 CCI comorbidity. Paclitaxel was more frequently prescribed than docetaxel.

**TABLE 1.**
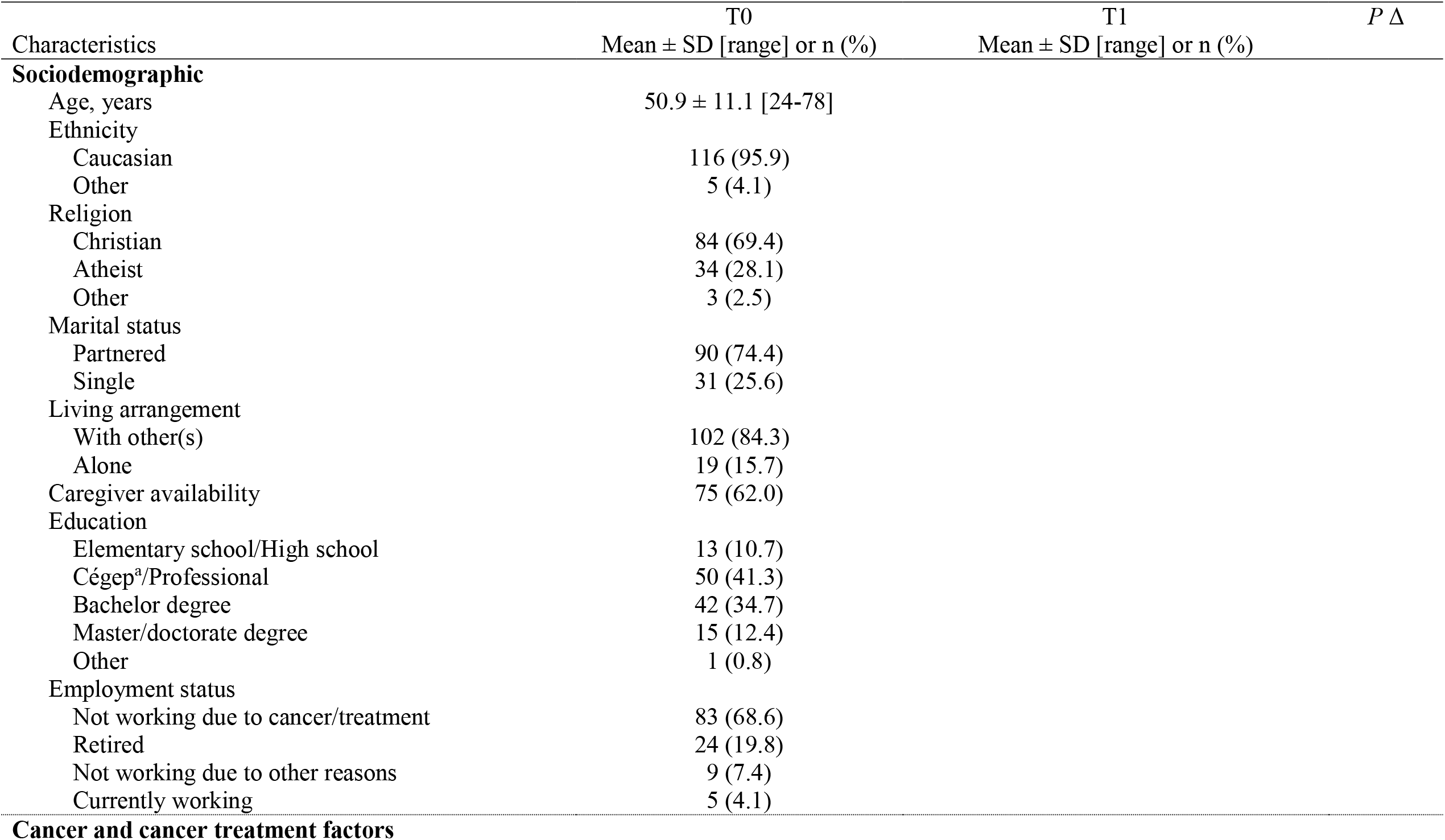
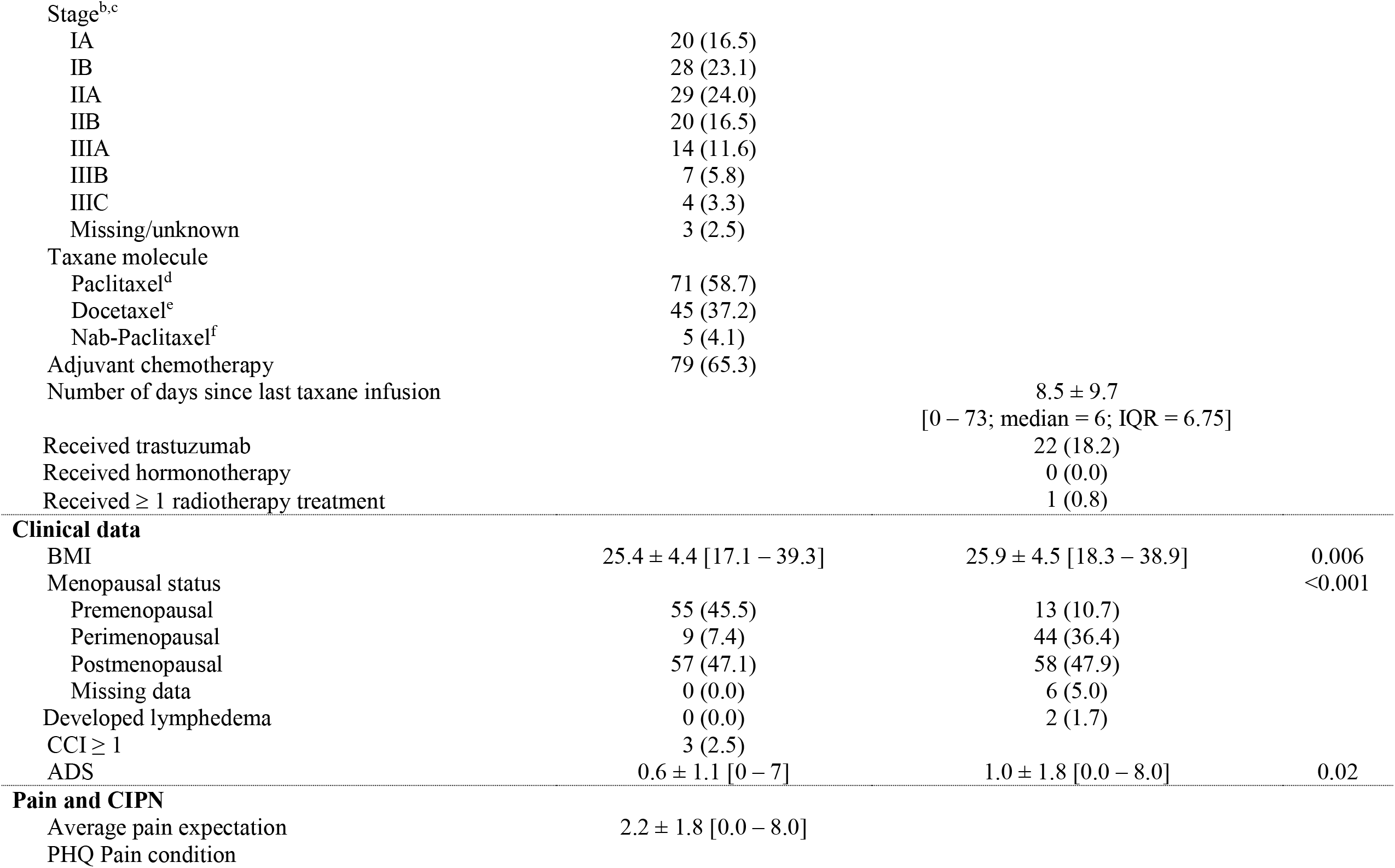
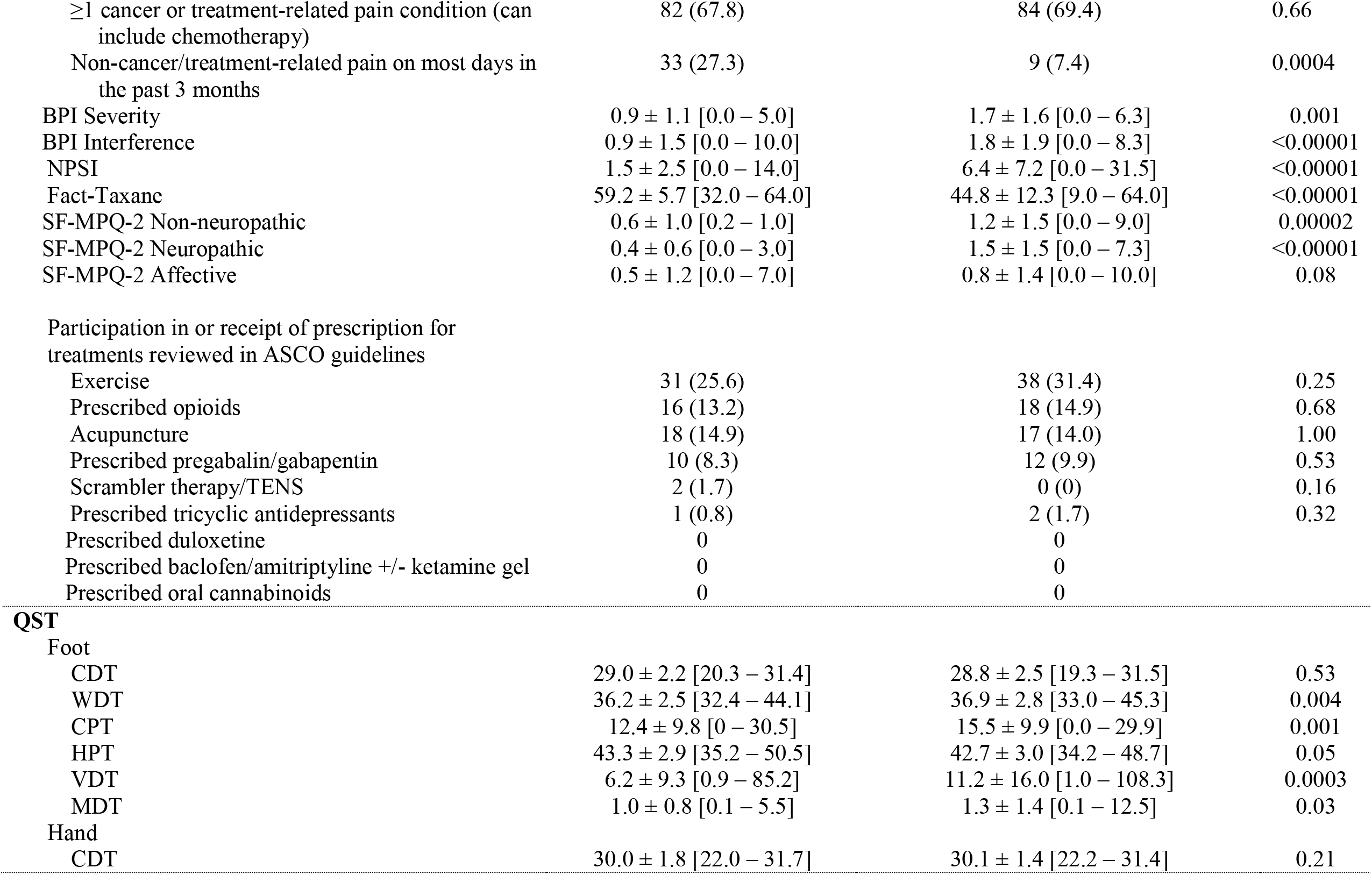
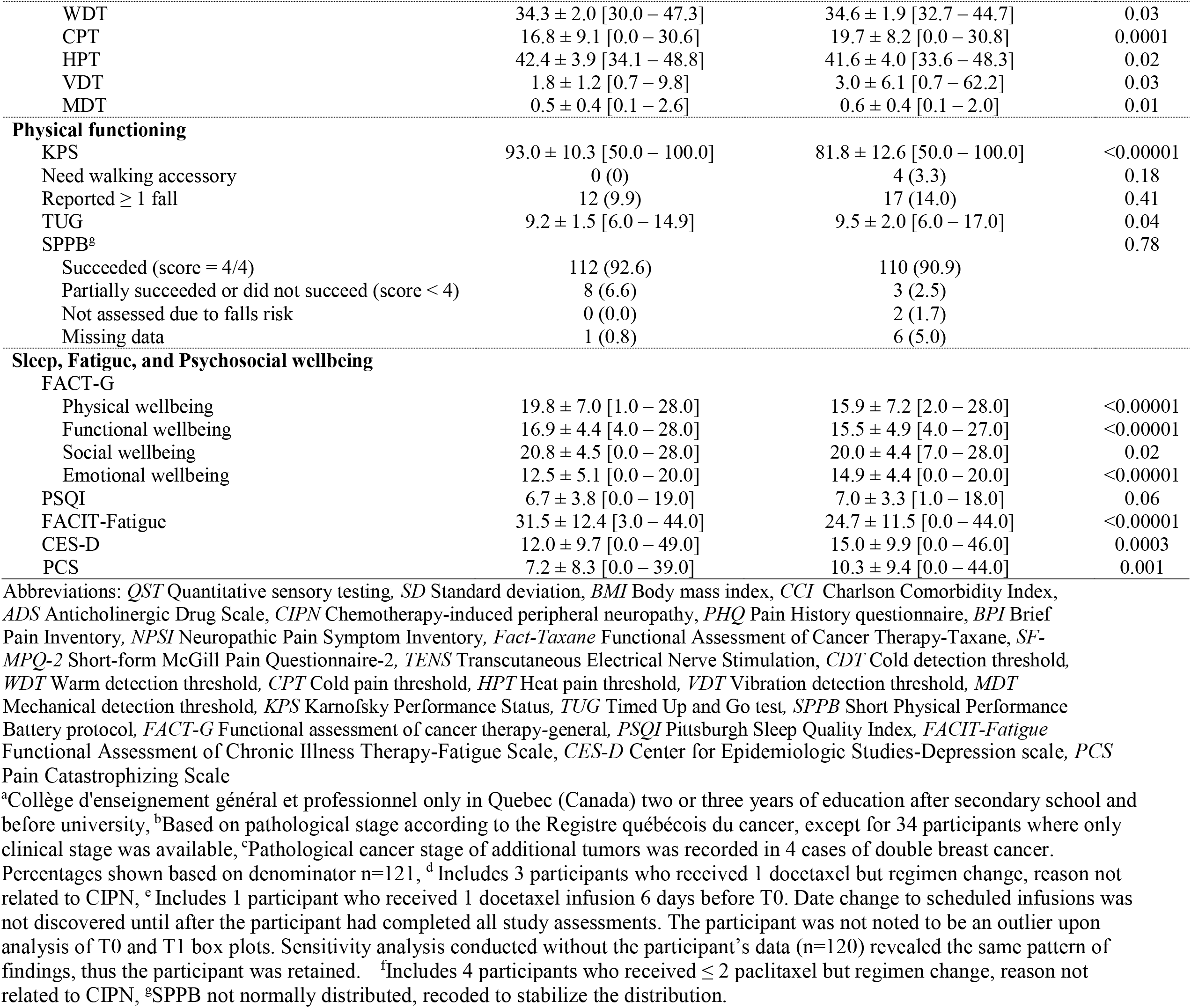
Sociodemographic and clinical characteristics, QST and physical assessment, self-report descriptive data, and T0 -T1 change.

With the exception of SF-MPQ-2 Affective, scores on all measures of pain and CIPN were significantly worse at T1 than T0. No participants were prescribed duloxetine. Frequency of pharmacological and nonpharmacological pain treatment frequency did not increase over time.

With the exception of foot and hand CDT, QST parameters were significantly different at T1 compared to T0, demonstrating nerve fiber functioning changes following chemotherapy. KPS was lower and TUG was higher at T1 than T0, suggesting worse observer-rated functioning and balance and mobility, respectively. Falls and SPPB did not change.

While PSQI did not change, FACIT-Fatigue and FACT-Physical, Functional, and Social wellbeing were worse at T1 than T0.

### Characterization of dose reduction (DR) and premature discontinuation (PD) outcome variable groups

Sixty-six (54.5%; G1) participants received taxane-as-prescribed. 46 (38.0%; G2) participants had neuropathy-related DR/PD (DR: n=37 [80.4%]; PD: n=5 [10.9%]; DR+PD: n=4 [8.7%]). This includes 3 women who had neuropathy and other symptoms. Nurses, pharmacists, and oncologists assessed patients using the National Cancer Institute Common Terminology Criteria for Adverse Events, a non-validated checklist including numbness, tingling, weakness, diminished or loss of sensation, and pain, or relied on patient self-report. There was no consistency in evaluation type. Nine (7.4%; G3) participants had DR/PD (DR n=5 [55.6%]; PD n=4 [44.4%]) due to other reasons, including febrile neutropenia (n=3), widespread muscle and joint pain and fatigue (n=1), dyspnea (n=1), cytopenia (n=1), disease progression (n=1), postphlebitic syndrome (n=1), and elevated alanine aminotransferase (n=1). G2 and G3 received 88.8%±11.1% (33.3%-98.7%) and 73.9%±32.0% (16.7%-98.2%) (G2 vs G3, p=0.20) of taxane dose, respectively.

### Bivariate analysis

Correlates retained for consideration for entry into the multivariable model were age, taxane molecule, T0 PCS and KPS, and T1 NPSI, Fact-Taxane, SF-MPQ-2 Neuropathic, foot and hand CPT, KPS and TUG (Table 2). T1 prescribed gabapentin/pregabalin, foot VDT, BPI Interference, and FACIT-Fatigue were not retained because group differences were only among G3 versus G1 or G2.

**TABLE 2.** Bivariate analysis comparing those who received taxane-as-prescribed (G1), those who had CIPN-related DR/PD (G2), and those who had DR/PD for other reasons (G3). Data are n(%) or Mean ± SD

|  | G1<br>(n = 66) | G2<br>(n = 46) | G3<br>(n = 9) | P | G1 | G2 | G3 | P |
| --- | --- | --- | --- | --- | --- | --- | --- | --- |
| Biopsychosocial factors |  |  |  |  |  |  |  |  |
| **Sociodemographic** |  |  |  |  |  |  |  |  |
| Age | 48.7 ± 10.1 | 53.8 ± 11.2 | 52.2 ± 15.2 | 0.05 | | | | |
| Caucasian | 63 (95.5) | 44 (95.7) | 9 (100.0) | 0.67 |  |  |  |  |
| Religion |  |  |  | 0.38 |  |  |  |  |
| Christian | 50 (75.8) | 30 (65.2) | 4 (44.4) |  |  |  |  |  |
| Atheist | 14 (21.2) | 15 (32.6) | 5 (55.6) |  |  |  |  |  |
| Other | 2 (3.0) | 1 (2.2) | 0 (0.0) |  |  |  |  |  |
| Marital status |  |  |  | 0.06 |  |  |  |  |
| Partnered | 48 (72.7) | 38 (82.6) | 4 (44.4) |  |  |  |  |  |
| Single | 18 (27.3) | 8 (17.4) | 5 (55.6) |  |  |  |  |  |
| Living arrangement |  |  |  | 0.48 |  |  |  |  |
| With other(s) | 54 (81.8) | 41 (89.1) | 7 (77.8) |  |  |  |  |  |
| Alone | 12 (18.2) | 5 (10.9) | 2 (22.2) |  |  |  |  |  |
| Caregiver availability | 41 (62.1) | 25 (54.3) | 6 (6.7) | 0.94 |  |  |  |  |
| Education |  |  |  | 0.88 |  |  |  |  |
| Elementary school/High school | 6 (9.1) | 5 (10.9) | 2 (22.2) |  |  |  |  |  |
| Cégep<sup>a</sup>/Professional | 29 (43.9) | 17 (37.0) | 4 (44.4) |  |  |  |  |  |
| Bachelor degree | 23 (34.8) | 17 (37.0) | 2 (22.2) |  |  |  |  |  |
| Master/doctorate degree | 7 (10.6) | 7 (15.2) | 1 (11.1) |  |  |  |  |  |
| Other | 1 (1.5) | 0 (0.0) | 0 (0.0) |  |  |  |  |  |
| Employment status |  |  |  | 0.25 |  |  |  |  |
| Not working due to cancer/treatment | 48 (72.7) | 30 (65.2) | 5 (55.6) |  |  |  |  |  |
| Retired | 9 (13.6) | 12 (26.1) | 3 (33.3) |  |  |  |  |  |
| Not working due to other reasons | 5 (7.6) | 4 (8.7) | 0 (0.0) |  |  |  |  |  |
| Currently working | 4 (6.1) | 0 (0.0) | 1 (11.1) |  |  |  |  |  |
| **Cancer and cancer treatment factors** |  |  |  |  |  |  |  |  |
| Cancer stage |  |  |  | 0.65 |  |  |  |  |
| IA, IB | 21 (31.8) | 22 (47.8) | 2 (22.2) |  |  |  |  |  |
| IIA, IIB | 27 (40.9) | 16 (34.8) | 5 (55.6) |  |  |  |  |  |
| IIIA, IIIB, IIIC | 16 (24.2) | 8 (17.4) | 2 (22.2) |  |  |  |  |  |
| Missing/unknown | 2 (3.0) | 0 (0) | 0 (0) |  |  |  |  |  |
| Taxane molecule |  |  |  | 0.0003 |  |  |  |  |
| Paclitaxel<sup>b</sup> | 34 (51.5) | 39 (84.8) | 3 (33.3) |  |  |  |  |  |
| Docetaxel | 32 (48.5) | 7 (15.2) | 6 (66.7) |  |  |  |  |  |
| Adjuvant chemotherapy | 43 (65.2) | 31 (67.4) | 5 (55.6) | 0.79 |  |  |  |  |
| Neoadjuvant chemotherapy | 23 (34.8) | 15 (32.6) | 4 (44.4) |  |  |  |  |  |
| Number of days since last taxane infusion |  |  |  |  | 7.5 ± 6.6 | 7.3 ± 7.6 | 20.9 ± 23.3 | 0.26<sup>c</sup> |
| Received Trastuzumab |  |  |  |  | 15 (22.7) | 7(15.2) | 0 (0.0) | 0.20 |
| **Clinical data** |  |  |  |  |  |  |  |  |
| BMI | 24.9 ± 4.4 | 26.2 ± 4.3 | 24.9 ± 4.9 | 0.32 | 25.5 ± 4.5 | 26.5 ± 4.5 | 25.2 ± 5.2 | 0.53 |
| Menopausal status |  |  |  | 0.32 |  |  |  | 0.08 |
| Premenopausal | 35 (53.0) | 16 (34.8) | 4 (44.4) |  | 11 (16.7) | 1 (2.2) | 1 (11.1) |  |
| Perimenopausal | 4 (6.1) | 5 (10.9) | 0 (0.0) |  | 25 (37.9) | 18 (39.1) | 2 (22.2) |  |
| Postmenopausal | 27 (40.9) | 25 (54.3) | 5 (55.6) |  | 27 (40.9) | 26 (56.5) | 5 (55.6) |  |
| Missing data | 0 (0.0) | 0 (0.0) | 0 (0.0) |  | 3 (4.5) | 1 (2.2) | 1 (11.1) |  |
| CCI ≥ 1 | 0 (0.0) | 3 (6.5) | 0 (0.0) | 0.08 |  |  |  |  |
| ADS | 0.5 ± 1.1 | 0.7 ± 1.3 | 0.8 ± 0.8 | 0.68 | 0.9 ± 1.8 | 1.0 ± 1.9 | 1.1 ± 2.3 | 0.95 |
| **Pain and CIPN** |  |  |  |  |  |  |  |  |
| Average pain expectation | 2.2 ± 1.7 | 2.5 ± 1.9 | 1.0 ± 1.6 | 0.09 |  |  |  |  |
| PHQ Pain condition |  |  |  |  |  |  |  |  |
| ≥1 cancer/cancer treatment-related pain condition (can include chemotherapy) | 41 (62.1) | 33 (71.7) | 8 (88.9) | 0.21 | 43 (65.2) | 33 (71.7) | 8 (88.9) | 0.29 |
| Non-cancer-related pain on<br>most days in the past 3<br>months | 16 (24.2) | 13 (28.3) | 4 (44.4) | 0.44 | 3 (4.5) | 5 (10.9) | 1 (11.1) | 0.40 |
| **BPI** |  |  |  |  |  |  |  |  |
| Severity | 0.9 ± 1.1 | 0.9 ± 1.0 | 1.6 ± 1.3 | 0.30 | 1.6 ± 1.7 | 1.7 ± 1.7 | 1.7 ± 1.4 | 0.98 |
| Interference | 0.8 ± 1.4 | 0.9 ± 1.7 | 1.5 ± 1.3 | 0.64 | 1.8 ± 2.1 | 2.0 ± 1.9 | 0.7 ± 0.8 | 0.01<sup>c</sup> |
| NPSI | 1.2 ± 2.3 | 1.7 ± 2.8 | 2.7 ± 1.8 | 0.32 | 4.6 ± 6.7 | 9.2 ± 7.0 | 4.3 ± 7.6 | 0.003 |
| FACT-Taxane subscale | 60.0 ± 5.8 | 58.3 ± 5.8 | 58.0 ± 4.0 | 0.28 | 48.7 ± 11.7 | 38.2 ± 10.6 | 54.3 ± 7.1 | 0.0001 |
| **SF-MPQ-2** |  |  |  |  |  |  |  |  |
| Non-neuropathic | 0.6 ± 1.0 | 0.6 ± 0.9 | 0.8 ± 0.9 | 0.88 | 1.0 ± 1.3 | 1.5 ± 1.7 | 0.7 ± 1.1 | 0.23 |
| Neuropathic | 0.3 ± 0.5 | 0.4 ± 0.7 | 0.5 ± 0.6 | 0.47 | 0.9 ± 1.3 | 2.5 ± 1.4 | 0.6 ± 1.0 | 0.0001 |
| Affective | 0.6 ± 1.4 | 0.4 ± 0.8 | 1.1 ± 1.4 | 0.33 | 0.5 ± 1.3 | 0.6 ± 1.3 | 0.9 ± 1.6 | 0.43 |
| **Pain treatments** |  |  |  |  |  |  |  |  |
| Exercise | 15 (22.7) | 15 (32.6) | 1 (11.1) | 0.29 | 22 (33.3) | 15 (32.6) | 1 (11.1) | 0.39 |
| Prescribed opioids | 8 (12.1) | 7 (15.2) | 1 (11.1) | 0.88 | 11 (16.7) | 5 (10.9) | 2 (22.2) | 0.59 |
| Acupuncture | 9 (13.6) | 8 (17.4) | 1 (11.1) | 0.81 | 8 (12.1) | 9 (20.0) | 0 (0) | 0.23 |
| Prescribed<br>pregabalin/gabapentin | 4 (6.1) | 5 (10.9) | 1 (11.1) | 0.63 | 5 (7.6) | 4 (8.7) | 3 (33.3) | 0.05<sup>d</sup> |
| **QST** |  |  |  |  |  |  |  |  |
| **Foot** |  |  |  |  |  |  |  |  |
| CDT | 28.9 ± 2.3 | 29.1 ± 2.2 | 29.3 ± 2.2 | 0.75 | 28.5 ± 2.8 | 29.1 ± 2.0 | 30.2 ± 1.3 | 0.13 |
| WDT | 36.1 ± 2.5 | 36.4 ± 2.5 | 36.4 ± 2.1 | 0.68 | 36.9 ± 2.8 | 37.0 ± 3.0 | 36.6 ± 2.5 | 0.87 |
| CPT | 11.3 ± 10.3 | 14.5 ± 9.3 | 12.3 ± 8.6 | 0.33 | 13.7 ± 10.0 | 18.9 ± 9.2 | 12.4 ± 7.3 | 0.02 |
| HPT | 43.2 ± 3.0 | 43.2 ± 2.8 | 44.0 ± 3.0 | 0.70 | 42.9 ± 3.0 | 42.3 ± 3.3 | 42.8 ± 1.8 | 0.56 |
| VDT | 6.5 ± 11.6 | 5.6 ± 5.6 | 6.2 ± 4.7 | 0.88 | 8.6 ± 11.9 | 16.0 ± 21.0 | 6.1 ± 5.4 | 0.04<sup>c</sup> |
| MDT | 1.0 ± 0.7 | 1.0 ± 0.9 | 0.9 ± 0.5 | 0.93 | 1.1 ± 0.9 | 1.5 ± 1.9 | 0.8 ± 0.5 | 0.21 |
| **Hand** |  |  |  |  |  |  |  |  |
| CDT | 30.1 ± 1.8 | 29.8 ± 1.8 | 29.4 ± 2.2 | 0.54 | 30.2 ± 1.1 | 30.0 ± 1.9 | 30.2 ± 1.4 | 0.79 |
| WDT | 34.1 ± 1.9 | 34.5 ± 2.3 | 34.1 ± 1.1 | 0.62 | 34.4 ± 2.0 | 34.8 ± 1.9 | 34.8 ± 1.7 | 0.64 |
| CPT | 15.1 ± 9.1 | 19.1 ± 8.5 | 17.0 ± 10.3 | 0.09 | 18.3 ± 8.1 | 22.3 ± 7.6 | 17.1 ± 9.4 | 0.03 |
| HPT | 42.6 ± 3.8 | 41.9 ± 4.0 | 43.2 ± 4.4 | 0.52 | 42.0 ± 3.9 | 40.8 ± 3.9 | 42.2 ± 4.3 | 0.28 |
| VDT | 1.7 ± 1.4 | 1.8 ± 0.9 | 1.7 ± 0.9 | 0.94 | 3.1 ± 7.7 | 3.0 ± 3.2 | 2.3 ± 1.7 | 0.94 |
| MDT | 0.5 ± 0.5 | 0.4 ± 0.4 | 0.5 ± 0.3 | 0.97 | 0.6 ± 0.4 | 0.6 ± 0.4 | 0.7 ± 0.6 | 0.62 |
| **Physical functioning** |  |  |  |  |  |  |  |  |
| KPS | 95.5 ± 7.4 | 90.4 ± 12.7 | 87.8 ± 10.9 | 0.02<sup>c</sup> | 85.3 ± 12.5 | 76.3 ± 11.4 | 83.3 ± 9.4 | 0.001 |
| Reported ≥ 1 fall | 5 (7.6) | 4 (8.7) | 3 (33.3) | 0.10 | 11 (16.7) | 5 (10.9) | 1 (11.1) | 0.64 |
| TUG | 9.1 ± 1.2 | 9.4 ± 1.8 | 9.6 ± 1.7 | 0.27 | 9.2 ± 1.7 | 10.1 ± 2.3 | 9.3 ± 2.5 | 0.04 |
| SPPB |  |  |  | 0.22 |  |  |  | 0.78 |
| Succeeded (score = 4/4) | 63 (95.5) | 41 (89.1) | 8 (88.9) |  | 62 (93.9) | 40 (87.0) | 8 (88.9) |  |
| Partially succeeded or did not succeed (score < 4) | 2 (3.0) | 5 (10.9) | 1 (11.1) |  | 1 (1.5) | 2 (4.3) | 0 (0.0) |  |
| Not assessed due to falls risk | 0 (0.0) | 0 (0.0) | 0 (0.0) |  | 1 (1.5) | 1 (2.2) | (0.0) |  |
| Missing data | 1 (1.5) | 0 (0.0) | 0 (0.0) |  | 2 (3.0) | 3 (6.5) | 1 (11.1) |  |
| **Psychosocial wellbeing** |  |  |  |  |  |  |  |  |
| FACT-G |  |  |  |  |  |  |  |  |
| Physical wellbeing | 20.6 ± 6.7 | 19.0 ± 7.6 | 18.3 ± 5.4 | 0.44 | 15.6 ± 7.3 | 15.5 ± 6.8 | 21.2 ± 8.2 | 0.18 |
| Functional wellbeing | 16.9 ± 4.7 | 16.9 ± 4.1 | 16.0 ± 2.5 | 0.88 | 15.5 ± 5.1 | 15.2 ± 4.7 | 18.7 ± 3.9 | 0.25 |
| Social wellbeing | 21.1 ± 4.3 | 21.1 ± 3.9 | 16.9 ± 8.8 | 0.08 | 19.9 ± 4.9 | 20.0 ± 3.9 | 21.7 ± 3.9 | 0.66 |
| Emotional wellbeing | 12.0 ± 5.6 | 13.2 ± 4.6 | 12.5 ± 4.4 | 0.53 | 14.9 ± 4.5 | 15.1 ± 4.2 | 13.2 ± 5.3 | 0.62 |
| PSQI | 6.6 ± 3.8 | 6.7 ± 3.9 | 8.0 ± 2.8 | 0.69 | 6.8 ± 3.2 | 7.7 ± 3.5 | 4.6 ± 2.4 | 0.07 |
| FACIT-fatigue | 31.9 ± 12.8 | 31.2 ± 12.3 | 30.3 ± 8.8 | 0.93 | 24.9 ± 12.2 | 22.6 ± 10.3 | 36.0 ± 5.4 | 0.03 |
| CES-D | 12.4 ± 10.4 | 11.2 ± 9.1 | 12.5 ± 5.2 | 0.81 | 16.1 ± 11.1 | 13.2 ± 8.3 | 15.5 ± 6.8 | 0.32<sup>c</sup> |
| PCS | 8.8 ± 9.9 | 4.9 ± 5.1 | 7.2 ± 6.5 | 0.05<sup>c</sup> | 11.6 ± 10.2 | 9.0 ± 8.6 | 6.0 ± 5.2 | 0.22 |

Abbreviations: ANOVA analysis of variance, *G1* Taxane as prescribed, *G2* neuropathy-related DR/PD, *G3* DR/PD due other reasons, *SD* Standard deviation, *BMI* Body mass index, *CCI* Charlson Comorbidity Index, *ADS* Anticholinergic Drug Scale, *CIPN* Chemotherapy-induced peripheral neuropathy, *PHQ* Pain History Questionnaire, *BPI* Brief Pain Inventory, *NPSI* Neuropathic Pain Symptom Inventory, *Fact-Taxane* Functional Assessment of Cancer Therapy-Taxane, *SF-MPQ-2* Short-form McGill Pain Questionnaire-2, *TENS* Transcutaneous Electrical Nerve Stimulation, *QST* Quantitative sensory testing, *CDT* Cold detection threshold, *WDT* Warm detection threshold, *CPT* Cold pain threshold, *HPT* Heat pain threshold, *VDT* Vibration detection threshold, *MDT* Mechanical detection threshold, *KPS* Karnofsky Performance Status, *TUG* Timed Up and Go test, *SPPB* Short Physical Performance Battery protocol, *FACT-G* Functional Assessment of Cancer Therapy-general, *PSQI* Pittsburgh Sleep Quality Index, *FACIT-Fatigue* Functional Assessment of Chronic Illness Therapy-Fatigue Scale, *CES-D* Center for Epidemiologic Studies-Depression scale, *PCS* Pain Catastrophizing Scale
<sup>a</sup>Collège d'enseignement général et professionnel only in Quebec (Canada) two or three years of education after secondary school and before university, <sup>b</sup>Nab-paclitaxel and paclitaxel are collapsed, <sup>c</sup>Homogeneity of variance assumption was violated, Welch F test was used, <sup>d</sup>Post-hoc test shows Group 3 received more Gabapentin/Pregabalin than Group 1 and 2, $p < 0.5$ .

T0 and T1 foot and hand CPT were highly correlated (*r*≥0.71, *p*<0.001). We retained both based on CIPN localization differences^61^. T1 NPSI and SF-MPQ-2 Neuropathic were also highly correlated (*r*=0.76, *p*<0.001). SF-MPQ-2 Neuropathic was retained over the NPSI due to larger G1 vs G2 effect size (Cohen’s *d*= -1.2; and -0.7, respectively).

### Logistic regression: Factors associated with neuropathy-related DR/PD

All variance inflation factors (VIF) were acceptable at ≤3.03^59^, suggesting moderate correlations. This is expected because of the repeated-measures nature of the data and does not require correction^62^. The model (Table 3) was statistically significant χ^2^(16)=88.64, *p*<0.001, explains 85% of the variance (Nagelkerke R^2^), and correctly classifies 94.4% of participants. Sample size is sufficient^63^. Paclitaxel (Odds Ratio [OR]=75.05, 95% Confidence Interval [CI], 2.56-2197.96), higher T1 SF-MPQ-2 Neuropathic pain (OR=10.77, 95% CI, 1.99-58.15) and hand CPT (OR=1.64, 95% CI, 1.05-2.56), and lower T0 PCS (0.72, 95% CI, 0.54-0.95) were associated with neuropathy-related DR/PD.

**TABLE 3.** Binomial logistic regression of factors associated with neuropathy-related DR/PD

| Predictors | B | S.E. | Wald | *p* | Odds Ratio | 95% CI for Odds Ratio |  |
| --- | --- | --- | --- | --- | --- | --- | --- |
|  |  |  |  |  |  | Lower | Upper |
| Taxane molecule (paclitaxel<sup>a</sup>) | 4.32 | 1.72 | 6.28 | 0.01 | 75.05 | 2.56 | 2197.96 |
| Age | 0.04 | 0.08 | 0.25 | 0.62 | 1.04 | 0.89 | 1.21 |
| T0 - KPS | -0.14 | 0.08 | 2.58 | 0.11 | 0.87 | 0.74 | 1.03 |
| T1 - KPS | -0.26 | 0.06 | 0.21 | 0.64 | 0.97 | 0.87 | 1.10 |
| T0 - QST |  |  |  |  |  |  |  |
| Foot CPT | 0.02 | 0.07 | 0.06 | 0.81 | 1.02 | 0.88 | 1.17 |
| Hand CPT | -0.29 | 0.15 | 3.67 | 0.06 | 0.75 | 0.55 | 1.01 |
| T1 - QST |  |  |  |  |  |  |  |
| Foot CPT | -0.10 | 0.09 | 1.00 | 0.32 | 0.92 | 0.77 | 1.10 |
| Hand CPT | 0.49 | 0.23 | 4.80 | 0.03 | 1.64 | 1.05 | 2.56 |
| T0 TUG | -0.39 | 0.54 | 0.51 | 0.47 | 0.68 | 0.23 | 1.97 |
| T1 TUG | 0.10 | 0.40 | 0.06 | 0.81 | 1.10 | 0.51 | 2.42 |
| T0 SF-MPQ-2 neuropathic scale | -0.32 | 1.37 | 0.06 | 0.81 | 0.72 | 0.05 | 10.48 |
| T1 SF-MPQ-2 neuropathic scale | 2.38 | 0.86 | 7.62 | 0.01 | 10.77 | 1.99 | 58.15 |
| T0 Fact-Taxane scale | -0.36 | 0.19 | 3.40 | 0.07 | 0.69 | 0.47 | 1.02 |
| T1 Fact-Taxane scale | -0.04 | 0.07 | 0.38 | 0.54 | 0.96 | 0.83 | 1.05 |
| T0 PCS Total score | -0.33 | 0.14 | 5.29 | 0.02 | 0.72 | 0.54 | 0.95 |
| T1 PCS Total score | -0.14 | 0.10 | 2.02 | 0.16 | 0.87 | 0.71 | 1.05 |

Abbreviations: *S.E.* Standard error, *KPS* Karnofsky performance status, *QST* Quantitative sensory testing, *CPT* cold pain threshold, *VDT* vibration detection threshold, *F-MPQ-2* Short-form McGill pain questionnaire-2, *Fact-Taxane* Functional assessment of cancer therapy-Taxane, *PCS* Pain catastrophizing scale
<sup>a</sup>Nab-paclitaxel and paclitaxel are collapsed; reference group is docetaxel

## DISCUSSION

This is the first study to examine biopsychosocial factors associated with CIPN-related DR/PD in women undergoing taxane-based chemotherapy for early-stage BC. 38% of women had CIPN-related DR/PD, the most common reason for DR/PD. Although our approach to determining whether DR/PD was CIPN-related was not based on a standardized clinical diagnosis, it reflects how CIPN is commonly assessed in clinical practice, thereby maximizing generalizability of findings to the community. Despite worse post-treatment CIPN and pain, no one was prescribed duloxetine, the only recommended agent for painful CIPN^4^. Prescriptions for other analgesics (e.g. gabapentin/pregabalin) and use of other treatments did not increase following chemotherapy. Contrary to assumptions that DR/PD exclusively reflects CIPN severity^10^, we found that the factors associated with DR/PD were biopsychosocial, including receipt of paclitaxel, greater post-treatment neuropathic pain and cold pain sensitivity in the hand, and lower pre-treatment pain catastrophizing. This study provides important information about the type of CIPN-related intolerance and functional nerve impairment associated with DR/PD decision-making to help refine guidelines.

Several findings are consistent with known risk factors for CIPN presence and severity. Consistent with the known dose limiting effects of paclitaxel^10,12,25^ and higher incidence of neurotoxicity with paclitaxel than docetaxel^64^, receipt of paclitaxel was associated with a greater likelihood of DR/PD than receipt of docetaxel.

The relationship of higher post-treatment neuropathic pain with CIPN-related DR/PD is not surprising given associations between self-report neuropathic pain and CIPN severity^36,71,72^. However, it is interesting that a similar relationship was not detected with FACT-Taxane scores. It is possible that patients report that symptoms are intolerable, or clinicians interpret them as such, when they reflect neuropathic pain, rather than non-painful neuropathy. It is also possible that the FACT-Taxane has insufficient sensitivity for CIPN-related DR/PD. Future research should compare the discriminant properties of CIPN and neuropathic pain measures across CIPN outcomes.

The relationship between high post-treatment hand CPT and CIPN-related DR/PD is consistent with research demonstrating that cold hyperalgesia in the upper limbs is an early marker of neurotoxicity in oxaliplatin treatment^33^. This important finding elucidates the type of impairment associated with CIPN-related DR/PD, suggesting that patients’ reports to clinicians of cold pain sensitivity in the upper limbs, which may reflect hyperalgesia or allodynia in the upper limbs, or clinician interpretation of such impairment, may play a role in CIPN-related DR/PD decision-making.

Interestingly, foot CPT was not associated with CIPN-related DR/PD. In fact, while lower limb disability, balance problems, and falls are associated with CIPN presence and severity^32,34,73,74^, in this study, these factors were not associated with CIPN-related DR/PD. It is possible that sensory abnormalities in the hands have a greater impact on daily life, or are interpreted as such, than sensory abnormalities in the feet, contributing to decision-making. Future research is needed to improve assessment and expand our understanding of lower limb impairment associated with CIPN-related DR/PD.

It is also interesting that QOL, depression, sleep and fatigue, which are associated with CIPN presence or severity^19–21^, were not associated with CIPN-related DR/PD, suggesting important variations in the factors important to each outcome. Some of this variation may be related to unknown factors guiding decision-making, which remains unclear, along with patients’ understanding of decisions, their implications, and the long-term impact of CIPN. We urgently need to improve our understanding to clarify decision-making guidance due to the unclear trajectory and reversibility of CIPN.

Most intriguingly, lower pre-treatment pain catastrophizing was associated with CIPN-related DR/PD. This may initially seem counterintuitive since higher catastrophizing is associated with greater severity of other types of cancer-related pain^75,76^. As DR/PD decisions rely on patient-clinician communication, symptom interpretation and communication models may be informative^71–73^. In particular, the Cancer Threat Interpretation Model^73^ proposes that bodily sensation appraisals play a role in patient-clinician communication, especially in uncertain health contexts. Pain catastrophizing is associated with cancer-related worry^80,81^. Therefore, high pre-treatment catastrophizing may reflect cancer-related worry, or a focus on worst-case possibilities in the face of survival uncertainty associated with DR/PD^82,83^, resulting in reluctance to disclose CIPN. In a qualitative study, fear of PD and resulting compromised treatment efficacy was a deterrent to CIPN disclosure^11^. Although this requires further investigation, taken together, symptom appraisal style may be critical to CIPN-related treatment decisions. Early and frequent CIPN assessment with valid and reliable tools are necessary to identify people who may underreport symptoms.

Limitations include the small sample size, preventing evaluation of DR independently from PD and the potential moderating role of dose received. Larger studies should stratify across these factors. Participants were mostly white, had few comorbidities, and were well-educated. Moreover, those who completed both assessments were younger than those who did not complete T1. While loss-to-attrition is generally higher among older adults in similar studies^78^, this may have generalizability implications. Finally, participants had early-stage BC. Factors important to CIPN-related DR/PD where the goals of care are palliative may be different^4^.

This study provides novel information about biopsychosocial factors associated with CIPN-related DR/PD. Intolerable CIPN and/or functional nerve impairment leading to DR/PD may reflect high neuropathic pain and cold pain sensitivity in the upper limbs, with communication of CIPN experience influenced by symptom appraisal. These data could help refine a recently proposed patient-centric decision-framework for CIPN-related treatment changes^14^ and contribute to clinical practice recommendations facilitating decision-making.

## Data Availability

Data available upon reasonable request

## ACKNOWLEDGMENTS

We thank the staff at Centre des maladies du sein Deschênes-Fabia, and Dominique St. Pierre, Marianne Vallée, and Katherine Bellavance for help with data collection, and members of the Cancer Pain and Supportive Care Research Lab for comments on earlier manuscript drafts. Most importantly, we thank the participants, whose time and efforts made this study possible.

## AUTHOR CONTRIBUTIONS

**Conception and design:** Lynn R. Gauthier, Lucia Gagliese, Jennifer Gewandter, Robert H Dworkin, Julie Lemieux, Josée Savard, Philip L. Jackson, Michèle Aubin, Sophie Lauzier

**Financial support:** Lynn R. Gauthier

**Administrative support:** Lynn R Gauthier, Maud Bouffard

**Provision of study materials or patients:** Lynn R Gauthier, Julie Lemieux, Philip L. Jackson, Anne Dionne

**Collection and assembly of data:** Lynn R. Gauthier, Lye-Ann Robichaud, Maud Bouffard, Frédérique Therrien, Sarah Béland, Marianne Bouvrette

**Data analysis and interpretation:** Lynn R Gauthier, Lye-Ann Robichaud

**Manuscript writing:** All authors

**Final approval of manuscript:** All authors

**Accountable for all aspects of the work:** All authors

## References

1. Wang Y-J, Chan Y-N, Jheng Y-W, et al: Chemotherapy-induced peripheral neuropathy in newly diagnosed breast cancer survivors treated with taxane: a prospective longitudinal study. Support Care Cancer 29:2959–2971, 2021

2. Eckhoff L, Knoop AS, Jensen MB, et al: Risk of docetaxel-induced peripheral neuropathy among 1,725 Danish patients with early stage breast cancer. Breast Cancer Res Treat 142:109–118, 2013

3. Hershman DL, Weimer LH, Wang A, et al: Association between patient reported outcomes and quantitative sensory tests for measuring long-term neurotoxicity in breast cancer survivors treated with adjuvant paclitaxel chemotherapy. Breast Cancer Res Treat 125:767–774, 2011

4. Loprinzi CL, Lacchetti C, Bleeker J, et al: Prevention and Management of Chemotherapy-Induced Peripheral Neuropathy in Survivors of Adult Cancers: ASCO Guideline Update. J Clin Oncol 38:3325–3348, 2020

5. Starobova H, Vetter I: Pathophysiology of chemotherapy-induced peripheral neuropathy. Front Mol Neurosci 10:174, 2017

6. Hershman DL, Till C, Wright JD, et al: Comorbidities and risk of chemotherapy-induced peripheral neuropathy among participants 65 years or older in southwest oncology group clinical trials. J Clin Oncol 34:3014–3022, 2016

7. Brewer JR, Morrison G, Dolan ME, et al: Chemotherapy-induced peripheral neuropathy: Current status and progress. Gynecol Oncol 140:176–183, 2016

8. Speck RM, Sammel MD, Farrar JT, et al: Impact of Chemotherapy-Induced Peripheral Neuropathy on Treatment Delivery in Nonmetastatic Breast Cancer. J Oncol Pract 9:e234–e240, 2013

9. Rosenbaek F, Holm HS, Hjelmborg JVB, et al: Effect of cryotherapy on dose of adjuvant paclitaxel in early-stage breast cancer. Support Care Cancer 28:3763–3769, 2020

10. Bhatnagar B, Gilmore S, Goloubeva O, et al: Chemotherapy dose reduction due to chemotherapy induced peripheral neuropathy in breast cancer patients receiving chemotherapy in the neoadjuvant or adjuvant settings: A single-center experience. Springerplus 3:366, 2014

11. Salgado TM, Quinn CS, Krumbach EK, et al: Reporting of paclitaxel-induced peripheral neuropathy symptoms to clinicians among women with breast cancer: a qualitative study. Support Care Cancer 28:4163–4172, 2020

12. Wong ML, Cooper BA, Paul SM, et al: Age-related differences in patient-reported and objective measures of chemotherapy-induced peripheral neuropathy among cancer survivors. Support Care Cancer 27:3905–3912, 2019

13. Nyrop KA, Deal AM, Reeder-Hayes KE, et al: Patient-reported and clinician-reported chemotherapy-induced peripheral neuropathy in patients with early breast cancer: Current clinical practice. Cancer 125:2945–2954, 2019

14. Hertz DL, Childs DS, Park SB, et al: Patient-centric decision framework for treatment alterations in patients with Chemotherapy-induced Peripheral Neuropathy (CIPN). Cancer Treat Rev 99:102241, 2021

15. Hertz DL, Kidwell KM, Vangipuram K, et al: Paclitaxel plasma concentration after the first infusion predicts treatment-limiting peripheral neuropathy. Clin Cancer Res 24:3602–3610, 2018

16. Porter LS, Keefe FJ: Psychosocial issues in cancer pain. Curr Pain Headache Reports 15:263–270, 2011

17. Syrjala KL, Chapko ME: Evidence for a biopsychosocial model of cancer treatment-related pain. Pain 61:69–79, 1995

18. Seretny M, Currie GL, Sena ES, et al: Incidence, prevalence, and predictors of chemotherapy-induced peripheral neuropathy: A systematic review and meta-analysis. Pain 155:2461–2470, 2014

19. Bao T, Basal C, Seluzicki C, et al: Long-term chemotherapy-induced peripheral neuropathy among breast cancer survivors: prevalence, risk factors, and fall risk. Breast Cancer Res Treat 159:327–333, 2016

20. Salehifar E, Janbabaei G, Hendouei N, et al: Comparison of the Efficacy and Safety of Pregabalin and Duloxetine in Taxane-Induced Sensory Neuropathy: A Randomized Controlled Trial. Clin Drug Investig 40:249–257, 2020

21. Simon NB, Danso MA, Alberico TA, et al: The prevalence and pattern of chemotherapy-induced peripheral neuropathy among women with breast cancer receiving care in a large community oncology practice. Qual Life Res 26:2763–2772, 2017

22. Tofthagen C, McAllister RD, Visovsky C: Peripheral Neuropathy Caused by Paclitaxel and Docetaxel: An Evaluation and Comparison of Symptoms. J Adv Pract Oncol 4:204–215, 2013

23. Gewandter JS, Fan L, Magnuson A, et al: Falls and functional impairments in cancer survivors with chemotherapy-induced peripheral neuropathy (CIPN): A University of Rochester CCOP study. Support Care Cancer 21:2059–2066, 2013

24. Kolb NA, Smith AG, Singleton JR, et al: The Association of Chemotherapy-Induced Peripheral Neuropathy Symptoms and the Risk of Falling. JAMA Neurol 73:860–866, 2016

25. Zanville NR, Nudelman KNH, Smith DJ, et al: Evaluating the impact of chemotherapy-induced peripheral neuropathy symptoms (CIPN-sx) on perceived ability to work in breast cancer survivors during the first year post-treatment. Support Care Cancer 24:4779–4789, 2016

26. Zhi WI, Chen P, Kwon A, et al: Chemotherapy-induced peripheral neuropathy (CIPN) in breast cancer survivors: a comparison of patient-reported outcomes and quantitative sensory testing. Breast Cancer Res Treat 178:587–595, 2019

27. Lee KM, Jung D, Hwang H, et al: Pre-treatment anxiety is associated with persistent chemotherapy-induced peripheral neuropathy in women treated with neoadjuvant chemotherapy for breast cancer. J Psychosom Res 108:14–19, 2018

28. Kleckner IR, Jusko TA, Culakova E, et al: Longitudinal study of inflammatory, behavioral, clinical, and psychosocial risk factors for chemotherapy-induced peripheral neuropathy. Breast Cancer Res Treat 189:521–532, 2021

29. Hausheer FH, Schilsky RL, Bain S, et al: Diagnosis, management, and evaluation of chemotherapy-induced peripheral neuropathy. Semin Oncol 33:15–49, 2006

30. Flatters SJL, Dougherty PM, Colvin LA: Clinical and preclinical perspectives on Chemotherapy-Induced Peripheral Neuropathy (CIPN): A narrative review. Br J Anaesth 119:737–749, 2017

31. Kim JH, Dougherty PM, Abdi S: Basic science and clinical management of painful and non-painful chemotherapy-related neuropathy. Gynecol Oncol 136:453–459, 2015

32. Cavaletti G, Frigeni B, Lanzani F, et al: Chemotherapy-Induced Peripheral Neurotoxicity assessment: A critical revision of the currently available tools. Eur J Cancer 46:479–494, 2010

33. Attal N, Bouhassira D, Gautron M, et al: Thermal hyperalgesia as a marker of oxaliplatin neurotoxicity: a prospective quantified sensory assessment study. Pain 144:245–252, 2009

34. Hammond EA, Pitz M, Lambert P, et al: Quantitative sensory profiles of upper extremity chemotherapy induced peripheral neuropathy: Are there differences in sensory profiles for neuropathic versus nociceptive pain? Can J Pain 3:169–177, 2019

35. Forsyth PA, Balmaceda C, Peterson K, et al: Prospective study of paclitaxel-induced peripheral neuropathy with quantitative sensory testing. J Neurooncol 35:47–53, 1997

36. NIH National Institute on Alcohol Abuse and Alcoholism: Drinking levels defined [Internet] [cited 2018 Feb 1] Available from: https://www.niaaa.nih.gov/alcohol-health/overview-alcohol-consumption/moderate-binge-drinking

37. Katzman R, Brown T, Fuld P, et al: Validation of a short Orientation-Memory-Concentration Test of cognitive impairment. Am J Psychiatry 140:734–739, 1983

38. Gewandter JS, Brell J, Cavaletti G, et al: Trial designs for chemotherapy-induced peripheral neuropathy prevention: ACTTION recommendations. Neurology 91:403–413, 2018

39. Ferland CE, Villemure C, Michon P-E, et al: Multicenter assessment of quantitative sensory testing (QST) for the detection of neuropathic-like pain responses using the topical capsaicin model. Can J Pain 2:266–279, 2018

40. Charlson ME, Pompei P, Ales KL, et al: A new method of classifying prognostic comorbidity in longitudinal studies: development and validation. J Chronic Dis 40:373–383, 1987

41. Carnahan RM, Lund BC, Perry PJ, et al: The Anticholinergic Drug Scale as a measure of drug-related anticholinergic burden: associations with serum anticholinergic activity. J Clin Pharmacol 46:1481–1486, 2006

42. Breivik H: The burden of central anticholinergic drugs increases pain and cognitive dysfunction. More knowledge about drug-interactions needed. Scand J Pain 17:186–188, 2017

43. Gagliese L, Jovellanos M, Zimmermann C, et al: Age-related patterns in adaptation to cancer pain: a mixed-method study. Pain Med 10:1050–1061, 2009

44. Cleeland CS, Ryan KM: Pain assessment: global use of the Brief Pain Inventory. Ann Acad Med Singapore 23:129–138, 1994

45. Dworkin RH, Turk DC, Revicki DA, et al: Development and initial validation of an expanded and revised version of the Short-form McGill Pain Questionnaire (SF-MPQ-2). Pain 144:35–42, 2009

46. Bouhassira D, Attal N, Fermanian J, et al: Development and validation of the Neuropathic Pain Symptom Inventory. Pain 108:248–257, 2004

47. Cella DF, Tulsky DS, Gray G, et al: The Functional Assessment of Cancer Therapy scale: development and validation of the general measure. J Clin Oncol 11:570–579, 1993

48. Buysse DJ, Reynolds CF, Monk TH, et al: The Pittsburgh Sleep Quality Index: a new instrument for psychiatric practice and research. Psychiatry Res 28:193–213, 1989

49. Yellen SB, Cella DF, Webster K, et al: Measuring fatigue and other anemia-related symptoms with the Functional Assessment of Cancer Therapy (FACT) measurement system. J Pain Symptom Manag 13:63–74, 1997

50. Radloff LS: The CES-D scale: A self-report depression scale for research in the general population. Appl Psychol Meas 1:385–401, 1977

51. Sullivan MJL, Bishop SR, Pivik J: The Pain Catastrophizing Scale: Development and validation. Psychol Assess 7:524–532, 1995

52. Geber C, Breimhorst M, Burbach B, et al: Pain in chemotherapy-induced neuropathy – More than neuropathic? Pain 154:2877–2887, 2013

53. Karnofsky DA, Burchenal JH: The clinical evaluation of chemotherapeutic agents in cancer, in Macleod CM (ed): Evaluation of chemotherapeutic agents. New York, Columbia University Press, 1949, pp 191–205.

54. Yates JW, Chalmer B, McKegney FP: Evaluation of patients with advanced cancer using the Karnofsky Performance Status. Cancer 45:2220–2224, 1980

55. Mathias S, Nayak U, Isaacs B: Balance in elderly patients: the “get up and go” test. Arch Phys Med Rehabil 67:387–389, 1986

56. Whitney JC, Lord SR, Close JCT: Streamlining assessment and intervention in a falls clinic using the Timed Up and Go Test and Physiological Profile Assessments. Age Ageing 34:567–571, 2005

57. Guralnik JM, Simonsick EM, Ferrucci L, et al: A short physical performance battery assessing lower extremity function: association with self-reported disability and prediction of mortality and nursing home admission. J Gerontol 49:M85–94, 1994

58. Little RJA: A Test of Missing Completely at Random for Multivariate Data with Missing Values. J Am Stat Assoc 83:1198–1202, 1988

59. Tabachnick BG, Fidell LS: Using Multivariate Statistics (ed 6). Boston, Pearson Education, 2013

60. Katz MH: Multivariable Analysis A Practical Guide for Clinicians. New York, Cambridge University Press, 1999

61. Miaskowski C, Mastick J, Paul SM, et al: Chemotherapy-Induced Neuropathy in Cancer Survivors. J Pain Symptom Manage 54:204–218, 2017

62. Kutner MH, Nachtsheim C, Neter J, et al: Applied linear statistical models (ed 5). New York, McGraw-Hill/Irwin, 2005

63. Peduzzi P, Concato J, Kemper E, et al: A simulation study of the number of events per variable in logistic regression analysis. J Clin Epidemiol 49:1373–1379, 1996

64. Argyriou AA, Kyritsis AP, Makatsoris T, et al: Chemotherapy-induced peripheral neuropathy in adults: A comprehensive update of the literature. Cancer Manag Res 6:135–147, 2014

65. Reyes-Gibby CC, Morrow PK, Buzdar A, et al: Chemotherapy-induced peripheral neuropathy as a predictor of neuropathic pain in breast cancer patients previously treated with paclitaxel. J Pain 10:1146–1150, 2009

66. Ventzel L, Jensen AB, Jensen AR, et al: Chemotherapy-induced pain and neuropathy: a prospective study in patients treated with adjuvant oxaliplatin or docetaxel. Pain 157:560–568, 2016

67. Bao T, Basal C, Seluzicki C, et al: Long-term chemotherapy-induced peripheral neuropathy among breast cancer survivors: prevalence, risk factors, and fall risk. Breast Cancer Res Treat 159:327–333, 2016

68. Gewandter JS, Fan L, Magnuson A, et al: Falls and functional impairments in cancer survivors with chemotherapy-induced peripheral neuropathy (CIPN): A University of Rochester CCOP study. Support Care Cancer 21:2059–2066, 2013

69. Jacobsen PB, Butler RW: Relation of cognitive coping and catastrophizing to acute pain and analgesic use following breast cancer surgery. J Behav Med 19:17–29, 1996

70. Azizoddin DR, Schreiber K, Beck MR, et al: Chronic pain severity, impact, and opioid use among patients with cancer: An analysis of biopsychosocial factors using the CHOIR learning health care system. Cancer 127:3254–3263, 2021

71. Craig KD: Social communication model of pain. Pain 156:1198–1199, 2015

72. Sullivan MJL: The communal coping model of pain catastrophising: Clinical and research implications. Can Psychol 53:32–41, 2012

73. Heathcote LC, Eccleston C: Pain and cancer survival: A cognitive-affective model of symptom appraisal and the uncertain threat of disease recurrence. Pain 158:1187–1191, 2017

74. Whitney CA, Dorfman CS, Shelby RA, et al: Reminders of cancer risk and pain catastrophizing: relationships with cancer worry and perceived risk in women with a first-degree relative with breast cancer. Fam Cancer 18:9–18, 2019

75. Bovbjerg D, Keefe F, Soo M, et al: Persistent breast pain in post-surgery breast cancer survivors and women with no history of breast surgery or cancer: associations with pain catastrophizing, perceived breast cancer risk, breast cancer worry, and emotional distress. Acta Oncol (Madr) 58:763–768, 2019

76. Robichaud M: Cognitive Behavior Therapy Targeting Intolerance of Uncertainty: Application to a Clinical Case of Generalized Anxiety Disorder. Cogn Behav Pract 20:251–263, 2013

77. Han PKJ, Gutheil C, Hutchinson RN, et al: Cause or Effect? The Role of Prognostic Uncertainty in the Fear of Cancer Recurrence. Front Psychol 11:626038, 2021

78. Leinert E, Singer S, Janni W, et al: The Impact of Age on Quality of Life in Breast Cancer Patients Receiving Adjuvant Chemotherapy: A Comparative Analysis From the Prospective Multicenter Randomized ADEBAR trial. Clin Breast Cancer 17:100–106, 2017

